# Genome wide association neural networks (GWANN) identify genes linked to family history of Alzheimer’s disease

**DOI:** 10.1101/2022.06.10.22276251

**Authors:** Upamanyu Ghose, William Sproviero, Laura Winchester, Najaf Amin, Taiyu Zhu, Danielle Newby, Brittany S. Ulm, Angeliki Papathanasiou, Liu Shi, Qiang Liu, Marco Fernandes, Cassandra Adams, Ashwag Albukhari, Majid Almansouri, Hani Choudhry, Cornelia van Duijn, Alejo Nevado-Holgado

## Abstract

Augmenting traditional genome wide association studies (GWAS) with advanced machine learning algorithms can allow the detection of novel signals in available cohorts. We introduce “Genome wide association neural networks (GWANN)”, a novel approach that uses neural networks (NNs) to perform a gene-level association study with family history of Alzheimer’s disease (AD). In UK Biobank, we defined cases (n=42,110) as those with AD or family history of AD and sampled an equal number of controls. The data was split into an 80:20 ratio of training and testing samples, and GWANN was trained on the former followed by identifying associated genes using its performance on the latter. Our method identified 18 genes to be associated with family history of AD. *APOE*, *BIN1, SORL1, ADAM10, APH1B,* and *SPI1* have also been identified by previous AD GWAS. *PCDH9, NGR3, ROR1, LINGO2, SMYD3,* and *LRRC7* were among the new genes that have been previously associated with neurofibrillary tangles or phosphorylated tau. Furthermore, there is evidence for differential transcriptomic or proteomic expression between AD and healthy brains for 11 of the 12 new genes. A series of post-hoc analyses resulted in a significantly enriched protein-protein interaction network (*P-value<1×10^−16^*), and enrichment of relevant disease and biological pathways such as focal adhesion (*P-value=1×10^−4^*), extracellular matrix organisation (*P-value=1×10^−4^*), Hippo signalling (*P-value=7×10^−4^*), Alzheimer’s disease (*P-value=3×10^−4^*), impaired cognition (*P-value=4×10^−3^*), and autism spectrum disorders (*P-value=1×10^−2^*). Applying NNs for GWAS illustrates their potential to complement existing algorithms and methods and enable the discovery of new associations without the need to expand existing cohorts.

## 1 Introduction

Alzheimer’s disease (AD) affects approximately 30 million people in the world, making it the most common form of dementia^1^. It is characterised by the build-up of Aβ and tau proteins in the brain, leading to neuronal death and impaired cognitive function^2^. In the last 10 years, genome wide association studies (GWAS) have revolutionised our understanding of the inherited basis of disease and they have been critical in identifying multiple risk loci and novel disease pathways associated with AD involving the microglia and lysosome^3^.

However, the classical GWAS analysis depends on sample size, and despite the number of SNPs identified until today, they still only explain a fraction of the heritability of the disease^4^. Gene-based methods have been developed to identify the joint effects of rare variants^5,6^ and common variants^7^, and gene-level analysis from GWAS summary data^8^. However, there are currently no methods to perform gene-based discovery using machine learning methods.

Along with the modern availability of large datasets^9–12^, to complement and enhance current GWAS methods, we propose to use an approach based on machine learning to shed light on more complex patterns in genomic mechanisms involving gene interactions and non-linear relationships. Machine learning methods, more particularly Neural Networks (NNs), have been instrumental in the advancement of multiple engineering industries due to their efficacy in analysing complex data patterns^13,14^, especially where large amounts of data are available. Compared to the success of classical GWAS, the successes of NN in gene discovery has been limited. NNs have recently been employed and tested on various complex traits and diseases including eye colour and schizophrenia^15^. Our aim was to develop NNs specialised to perform a gene-level association study using SNP data available in the UK Biobank (UKB)^16^. Our method is gene-based and considers groups of SNPs within and around each gene in the genome to establish the association of the gene with the phenotype of interest. In this paper we demonstrate the application of our new GWANN method as a complementary method to existing GWAS methods, to identify associations with familial history of Alzheimer’s disease/dementia, a proxy that has been successfully used to identify new genes for AD in the UKB^17,18^. We present the genetic associations to family history of AD found by the method, and systematically support the results with post-hoc enrichment analyses using transcriptomic data from post-mortem AD brains, biological pathways and gene ontologies, protein-protein interaction (PPI) data, disease and trait gene sets, and data about target tractability for drug development.

### 2 Results

#### 2.1 Identification of genes related to family history of AD using GWANN

We applied GWANN to 42,110 cases of AD family history, and an equal number of controls. 80% (n=67,376) of the data was used to train the NNs and 20% (n=16,844) was used as a held-out test set. The analysis was run at a gene-level, where the SNPs within a gene and in the flanking 2500 bp region were mapped to the gene. Each gene was divided into windows containing a maximum of 50 SNPs, and a separate NN (Figure 1) was trained for each window. A total of 70,848 gene windows were tested. In addition to the SNPs, age (field 21003), sex (field 31), the first six genetic principal components (PCs) obtained from UKB variables (field 22009), and education qualification (field 6138) were used as covariates. Education qualification was transformed into years of education using the International Standard Classification of Education (ISCED) encoding. Since NNs are inherently stochastic^13^, for each window, the method was run 16 times with different random seeds to get a stable aggregate performance metric on the held-out test set, and to then determine the level of statistical significance of this metric being significantly above chance predictions of family history of AD. The aggregate metric was compared to a null distribution obtained from simulated data generated using the ‘dummy’ method of PLINK 2.0^19^ to obtain a P-value. An empirical threshold, θ*_1_ = 1×10^−25^*, was determined such that 95% of the gene windows with *P-value <* θ*_1_* would also satisfy *P-value < 7.06×10^−7^—* the Bonferroni corrected genome wide significance threshold—if the method were repeated another 16 times with different random seeds (see STAR Methods for further details). This was to ensure that only the most confident hits were reported as significant associations. The negative log loss (NLL) of the NNs were used as the test metrics to evaluate significance, and for all post-hoc analyses. If multiple windows of a gene were significant, the window with the best test metric was selected.

**Figure 1.**
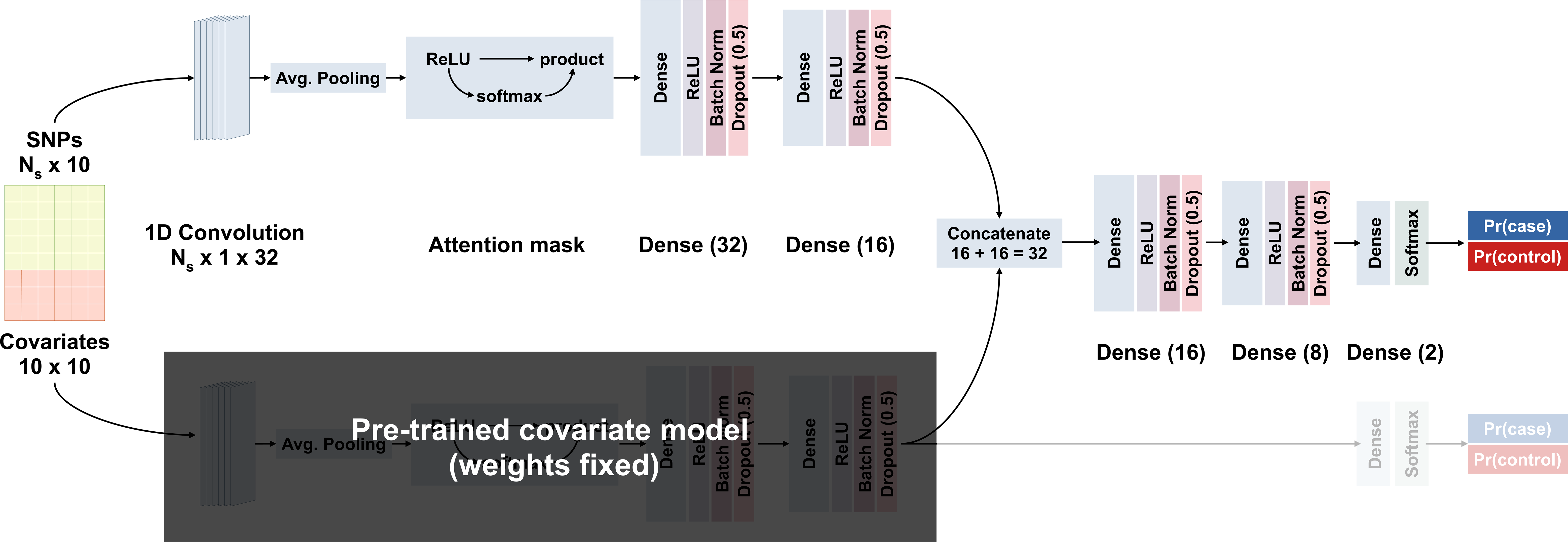
NN architecture used in the GWANN method. The top-left branch generates a 1D encoding from the SNPs input (green), while the bottom-left branch does so for the covariate input (red). The right trunk merges the encodings of both branches to output whether the input belongs to cases (blue output) or controls (red output).

32 genes passed the empirical significance threshold before pruning for linkage disequilibrium (LD). After identifying LD blocks (genes with r^2^ ≥ 0.8) among these genes, we retained the gene with the best test metric within a block as the hit gene. This resulted in narrowing down to 18 associated genes (Figure 2, Table 1). Among these hits, *APOE, BIN1, ADAM10, SORL1, SPI1,* and *APH1B* have been previously associated with AD by large GWAS^3^ (Figure 3). In addition to these AD associated genes, *LINGO2, LRRC7, NRG3, PCDH9, ROR1,* and *SMYD3* have been previously identified via SNP x SNP interaction studies to be associated with phosphorylated tau^3^. Six genes, *SYNPO, SRGAP2B, PALD1, AKR1C6P, HSP90AB4P,* and *RPS6KC1*, had no evidence for previous GWAS association with AD or AD-related traits. To further understand the 12 new GWANN hits, we obtained information about them from the Agora AD knowledge portal (https://agora.adknowledgeportal.org) (Figure 3). Besides *PCDH9* and *AKR1C6P*, all hits had evidence for differential transcriptomic or proteomic expression between post-mortem AD and healthy brains. RNAseq levels of *PCDH9* had evidence of association with clinical consensus diagnosis of cognitive status at time of death (COGDX). *AKR1E2,* a gene 23 kbp upstream of *AKR1C6P*, was nominally significant (*P-value = 7.43×10^−6^*) in the GWANN analysis, and also has evidence for association with phosphorylated tau in previous genome-wide interaction analysis^3^, and differential transcriptomic expression between AD and healthy brains (https://agora.adknowledgeportal.org). We also performed a GWAS using PLINK 2.0^19^ on the same data that was used for GWANN (TradGWAS). After LD pruning, TradGWAS identified *APOE* and *SORLI* as significant genes (*P-value < 5×10^−8^*), both of which were identified by GWANN. When compared with the genes identified by the largest AD GWAS run in the European population (EADB GWAS)^12^, GWANN had an overlap of five genes (*APOE, SORL1, APH1B, BIN1, SPI1)* and TradGWAS had an overlap of two genes (*APOE, SORL1*) (Figure 3c). We also looked at the overlap with the EADB GWAS hit genes using the top 100 genes from GWANN and TradGWAS (Figure 3d). This showed an overlap of 7 genes (*APOE, SORL1, BIN1, ABCA7, BCKDK, UFC1, CR1)* between the EADB GWAS and TradGWAS, and 7 genes (*APOE, SORL1, BIN1, ABCA7, APH1B, SPI1, CTSH)* between the EADB GWAS and GWANN. If an intergenic hit SNP in the EADB GWAS was not deterministically mapped to either the upstream or downstream gene, both were considered when calculating the overlap. The EADB GWAS reported 89 hit loci, but since we calculated the overlap on a gene level, we used the 84 unique genes that these loci were mapped to and added *APOE* to the list of hits.

**Figure 2:**
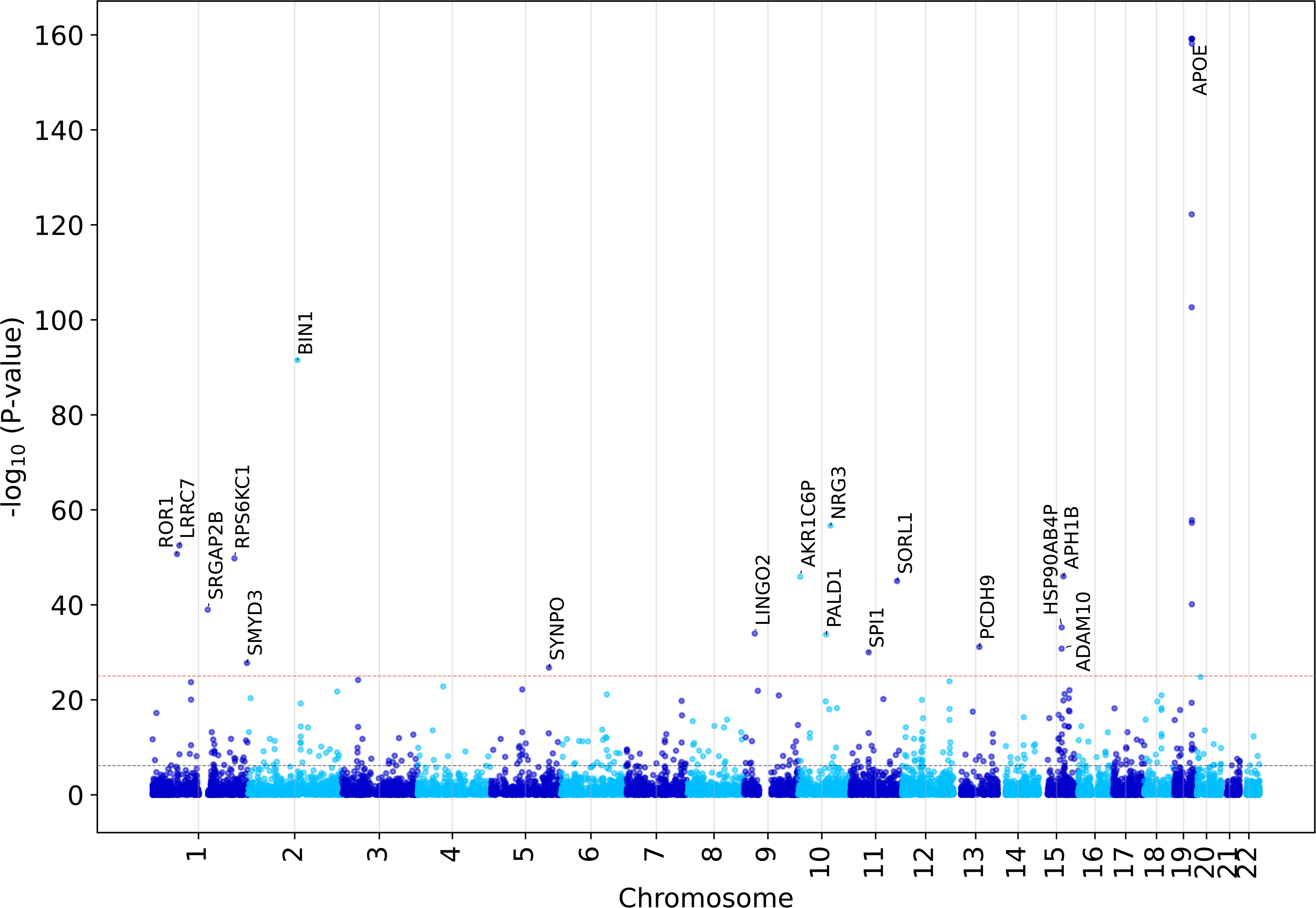
Manhattan plot after running GWANN on family history of AD. Significant hits identified at an empirically defined P-value threshold of P-value < 1×10^−25^ (red line). After calculating the LD between significant genes, the gene with the best negative log loss within an LD block was identified as the hit gene. The P-values lower than 6.95×10^−159^ have been cropped to a value of 6.95×10^−160^. The black line marks the Bonferroni corrected threshold for the number of gene windows that were tested, P-value = 7.06×10^−7^.

**Figure 3:**
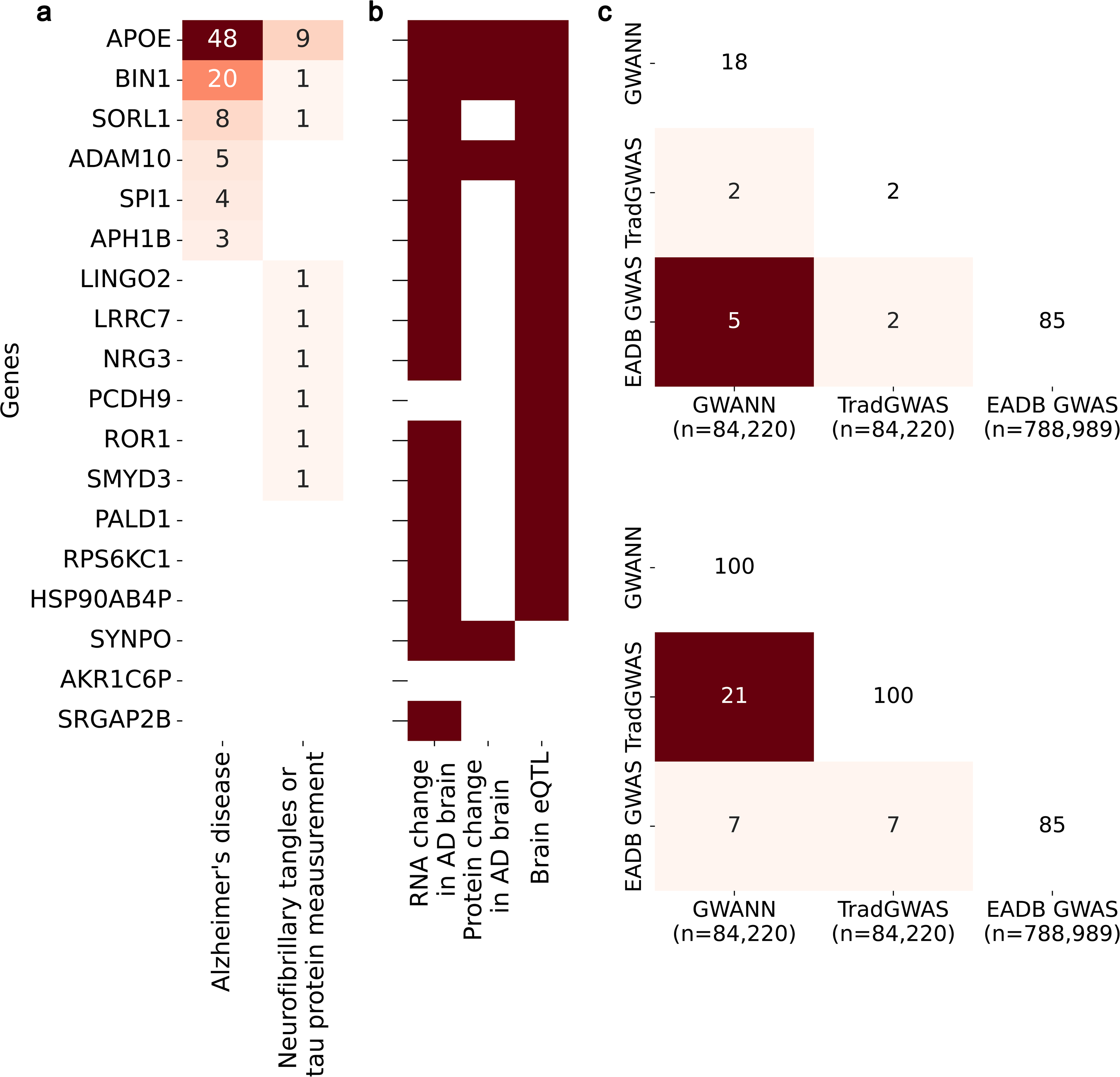
Overlap of GWANN hits with previous studies. (a) Heatmap showing the presence of significant evidence for the terms on the x-axis for the GWANN hits. (b) Heatmap showing the count of previous GWAS where the GWANN hits were identified to be associated with the phenotypes on the x-axis. (c) Heatmap showing the overlap between GWANN hits (GWANN), a GWAS run using PLINK 2.0 on the same data as GWANN (TradGWAS), and the largest European AD GWAS (EADB GWAS)^12^. (d) Similar heatmap to (c) but instead of using the GWANN and TradGWAS hits, the top 100 genes from both methods were considered for the overlap with the EADB GWAS hit genes. The sample size of each method is mentioned in the x-axis of the heatmaps, and the diagonals show the number of genes of each method considered while calculating the overlap.

**Table 1.**
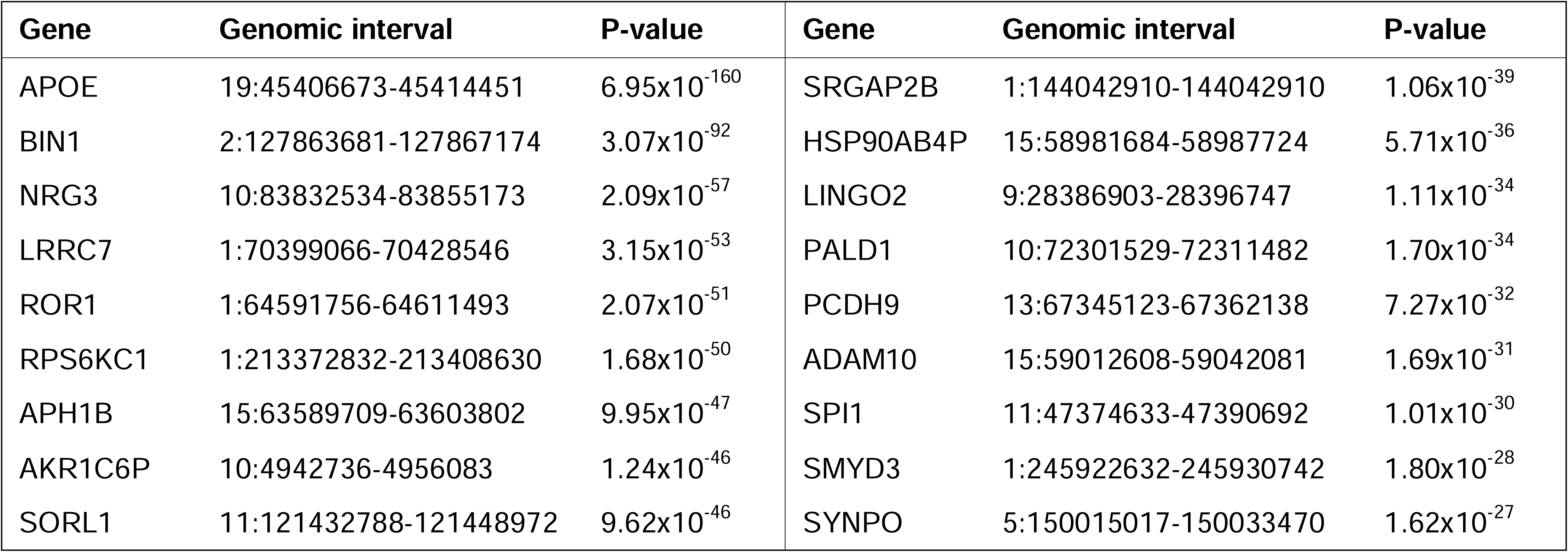
GWANN hit genes associated with family history of AD. P-values lower than 6.95×10^−159^ have been cropped to a value of 6.95×10^−160^.

#### 2.2 Enriched biological pathways, GO terms, diseases, and PPI network

Gene set enrichment analysis (GSEA)^20^ was applied to the GWANN summary test metrics for all genes to identify enriched pathways in Reactome, Wiki, Kyoto Encyclopaedia of Genes and Genomes (KEGG), and Gene Ontology (GO) gene sets obtained from MSigDB^21^ (Figure 4a-4d, Supplementary Table 2). GSEA calculates the normalised enrichment score (NES) based on the test metric of all genes analysed using a Kolmogorov-Smirnov-like test^20^. Hence, some pathways had a significant NES due to the cumulative contribution of genes that were nominally significant but not among the list of 18 GWANN hits. The enriched pathways with the GWANN hits present in the GSEA leading edge were extracellular matrix organization (*P-value = 1.04×10^−4^*), signalling by receptor tyrosine kinases (*P-value = 7.64×10^−4^*), axon guidance (*P-value = 2.23×10^−3^*), diseases of signal transduction by growth factor receptors and second messengers (*P-value = 1.11×10^−2^*), and ERBB signalling (*P-value = 2.29×10^−2^*). The most enriched GO terms with GWANN hits in the GSEA leading edge were regulation of neuron projection development (*P-value = 7.03×10^−8^*), glutamatergic synapse (*P-value = 2.64×10^−6^*), synapse organisation (*P-value = 5.66×10^−6^*), distal axon (*P-value = 7.77×10^−5^*), and regulation of synapse structure or activity (*P-value = 7.72×10^−4^*).

**Figure 4:**
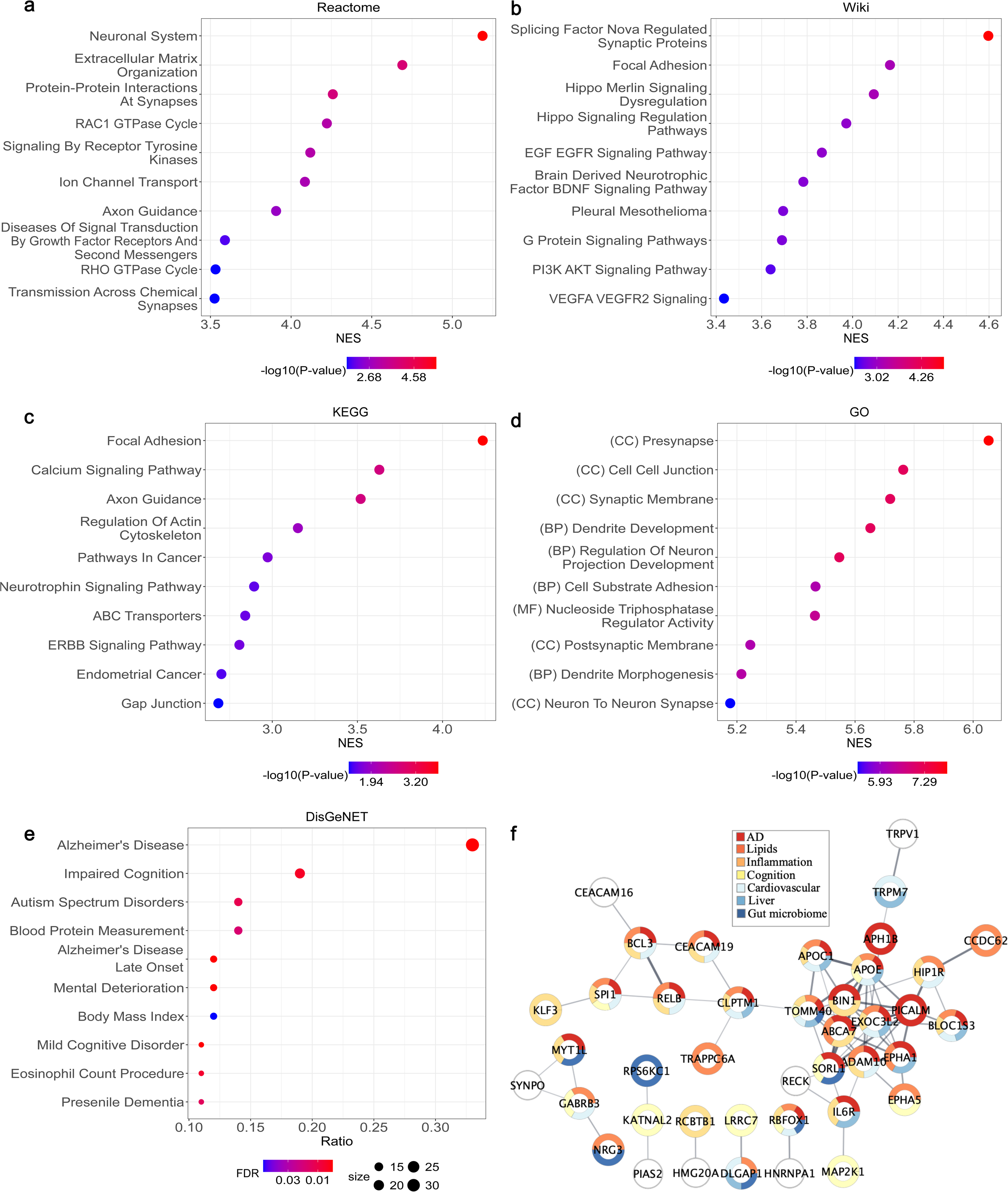
Post-hoc enrichment analysis after GWANN analysis. (a-d) Gene set enrichment analysis for Reactome (a), Wiki (b), KEGG (c), and GO (d) using GWANN summary metrics. (e-f) Genes were ranked according to the metric *1 – NLL_NN,_* where *NLL_NN_* was the negative log loss of the neural network for a given gene. (e) Disease and trait enrichment using the top 100 genes. (f) Enriched protein-protein interaction network (*P-value < 1 x 10^−16^*) for the top 100 genes. The colours within the nodes highlight the trait categories enriched by the protein encoded by the gene.

Disease and trait enrichment was performed using the DisGeNET^22^. The enrichment analysis requires a list of genes to perform an over representation analysis for all diseases and traits in the database. We used the top 100 genes (without LD pruning) ranked by the test metric for this analysis and filtered out diseases and traits with more than 5000 genes mapped to them (Figure 4e, Supplementary Table 3). Some of the most enriched traits with the largest number of overlapping genes were Alzheimer’s disease (*FDR = 2.56×10^−4^*), impaired cognition (*FDR = 4.23×10^−3^*), autism spectrum disorders (*FDR = 1.19×10^−2^*), blood protein measurement (*FDR = 1.82×10^−2^*), and mental deterioration (*FDR = 3.50×10^−4^*).

Using the same set of genes as used for the disease and trait enrichment, we generated a PPI network using STRING^23^ (Figure 4f). Some of the gene symbols were not recognised by the STRING protein database, leaving a set of 88 genes that were accepted. The resultant PPI network was significantly enriched with 72 edges (*P-value < 1×10^−16^*). Given a network of 88 proteins, the expected number of edges for a set of randomly selected proteins is 21, thereby rendering the GWANN PPI network to have significantly more connections than an equivalent network of random proteins. The nodes in the network enriched multiple gene sets in the experimental factor ontology (EFO), broadly grouped (Supplementary Table 4) into AD-related traits, lipid and lipoprotein measurements, inflammation markers, cognition, cardiovascular diseases, liver enzyme measurements, and gut microbiome measurement. The network in Figure 4f shows each node (protein) coloured by the group of EFO traits it enriched.

We also performed the same enrichments for TradGWAS (Supplementary Figure 1). Pathways related to ERBB signalling and calcium signalling overlapped with GWANN. The other top enriched pathways were mainly related to cholesterol, lipids, and lipoproteins (Supplementary Figure 1a-d). The PPI network showed similar levels of enrichment to the GWANN PPI network (*P-value < 1×10^−16^*, Supplementary Figure 1e).

#### 2.3 Enrichment of transcriptomic data from AD post-mortem brains using GWANN hits

We identified two previous studies that looked at differential transcriptomic expression between the brains of post-mortem AD cases and healthy controls. The first study, by Patel et. al.^24^. reported the results of a meta-analysis of DEGs between AD cases and controls in the cerebellum, frontal lobe, parietal lobe, and temporal lobe. We used the DEGs identified by them when comparing AD vs controls, and non-AD mental disorders vs controls. We also performed the enrichment for DEGs unique to AD and no other mental disorder. The second study, by Patel et. al.^25^ listed DEGs between (i) asymptomatic AD cases vs controls, (ii) symptomatic AD cases vs controls, and (iii) symptomatic vs asymptomatic AD cases in the cerebellum, entorhinal cortex, frontal lobe, and temporal lobe. We applied the GSEA algorithm to identify the level of enrichment of the DEG sets. Table 2 contains the adjusted P-value of enrichment of the DEG sets for each condition and in each brain region.

**Table 2.**
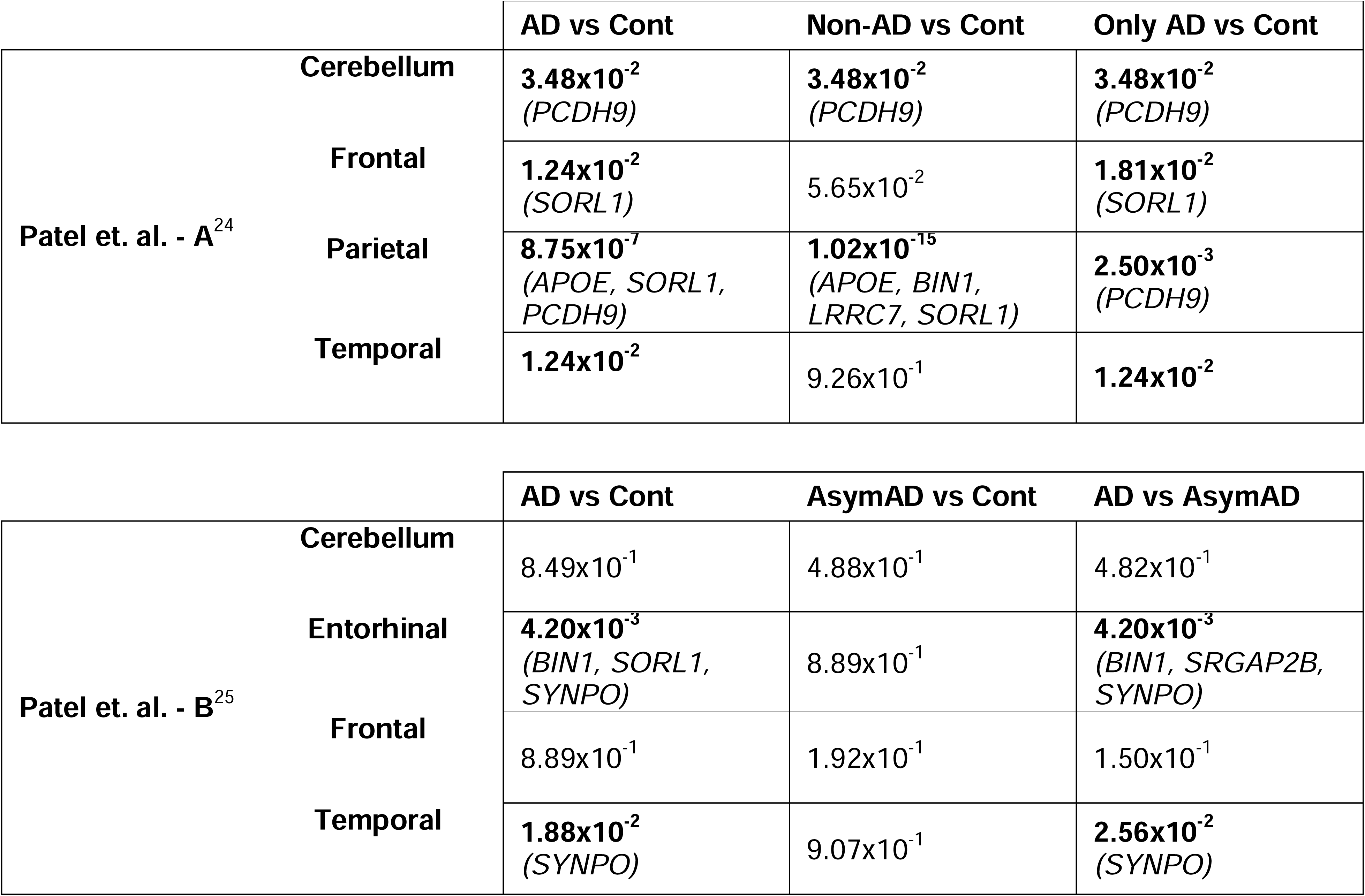
Enrichment of DEGs identified in two transcriptomic studies on AD brains. The genes mentioned in parentheses were the GWANN hits in the leading edge of the GSEA for each condition and brain region.

In the first study (Patel at. al. – A), DEG sets in all brain regions were enriched for AD vs controls and ‘Only AD’ vs controls, and the cerebellum and parietal lobes were enriched for ‘non-AD’ vs controls. The GWANN hits in the leading edge of the GSEA for the different enriched conditions and brain regions were *PCDH9, APOE, SORL1* and *LRRC7*. Despite the temporal lobe being enriched, none of the GWANN hits were in the leading edge. *UFC1*, a new hit identified in the EADB GWAS and a nominal hit in our analysis (*P-value = 2.4×10^−11^*), along with *MAP2K1*, another nominal GWANN hit (*P-value = 6.11×10^−22^*) were in the leading edge for the temporal lobe. Additionally, there was no difference between the DEGs for AD vs controls and ‘Only AD’ vs controls in the temporal lobe, thereby producing identical enrichment. There was no enrichment for the asymptomatic AD vs controls condition in the second study (Patel at. al. – B), and the entorhinal cortex and temporal lobes were enriched for symptomatic AD vs controls, and symptomatic AD vs asymptomatic AD. The GWANN hits in the leading edge were *BIN1, SORL1, SYNPO* and *SRGAP2B*.

The same enrichments were also performed for TradGWAS (GWAS on same data as GWANN) using PLINK 2.0 (Supplementary Table 7). The brain regions and conditions enriched were the same as that for GWANN.

#### 2.4 Potential of GWANN targets for AD drug discovery

To assess the tractability of the GWANN hits for aiding drug discovery, we used TargetDB^26^ to score them based on information collected from literature, and knowledge about their chemistry, biology, structure, and genetics. *ADAM10, APOE, SMYD3, BIN1, SORL1,* and *ROR1* were reported to be tractable; and *SPI1, LRRC7, APH1B, NRG3,* and *PCDH9* were reported as challenging, but tractable (Supplementary Table 5). Amongst the tractable genes, *ROR1* has a drug, Cirmtuzumab, associated with it which is currently under clinical trials for different cancers and neoplasms^27^.

### 3 Discussion

We developed GWANN and applied it to identify genes associated with family history of AD using data from the UKB. In doing so, we were able to identify 18 genes significantly associated with the phenotype. The post-hoc enrichment analyses showed enriched biological and disease pathways relevant to AD and neurodegeneration. Several GWANN hits were also identified as tractable drug targets.

#### 3.1 Role of hit genes and enriched biological pathways in neurodegeneration and AD

While some of the GWANN hits have not been identified in previous GWAS, many of them, or their associated biological pathways, have been linked to AD or neurodegeneration. For brevity, the six well-known AD genes identified by GWANN have not been discussed here. In the following paragraphs, the pathways and gene ontologies mentioned in parentheses were enriched and contains the gene being discussed.

*LINGO2* has been linked to promoting lysosomal degradation of amyloid-β protein precursor, thereby having a protective effect against AD^28^. A similar finding has been reported in a previous differential gene expression analysis in the CA1 and CA3 brain regions of the hippocampus, where they found higher expressions of *LINGO2* in healthy controls as compared to AD patients^29^. In our post-hoc enrichment analyses, *LINGO2* contributed to the enrichment of biological processes involved with synapse organisation (GO:0050808), and synapse activity and structure (GO:0050803). This could explain its importance in maintaining a healthy synapse by facilitating the clearance of amyloid-β, as identified by the previously mentioned studies. Additionally, although *LINGO2* has not been identified as a genome wide significant gene for AD, it has been identified to be nominally significant in previous GWAS on (i) non-hypertensive AD cases vs controls^30^, and (ii) brain region atrophy (entorhinal cortex thickness, hippocampus volume, ventricular volume) derived from MRI data^31^. Another GWANN hit, *SYNPO,* has demonstrated involvement in synaptic plasticity^32^. It has been previously shown to have a role in facilitating the autophagic clearance of p-Ser262 microtubule-associated protein tau (*MAPT*)^33^. Our post-hoc enrichment analyses highlighted its localisation to the actin cytoskeleton (GO:0015629). The role of the actin cytoskeleton in facilitating autophagy^34^ and the involvement of *SYNPO* with the organisation (GO:0007015) and binding (GO:0003779) of this cellular component could explain how it aids in the clearance of phosphorylated tau. Furthermore, it was also identified to be significantly downregulated in patients with dementia of Lewy bodies and Parkinson’s disease dementia, suggesting its role in other neurodegenerative disorders with similar pathology to AD^35^. Along with *SYNPO*, *ROR1* is among the new GWANN hits that also contributes to the maintenance of healthy synapses through its involvement with the cytoskeleton. It encodes a receptor tyrosine kinase (GO:0004713) that is associated with the actin cytoskeleton (GO:0015629). Actin filaments help in maintaining the integrity of the neuronal cytoskeleton, and past studies have implicated the role of amyloid-β in disrupting the cytoskeleton. The overexpression of *ROR1* was shown to stop cytoskeleton degradation *in-vitro*, even in the presence of amyloid-β by preserving the actin network^36^. Epigenetically, *ROR1* was also identified as a gene with differential hydroxymethylation between late-onset AD patients and healthy controls. Along with six other genes, it was shown to be correlated with MMSE and MoCA scores of subjects^37^.

*NRG3,* along with some of the well-known AD hits—*APOE, BIN1, APH1B, ADAM10*—, is part of the enriched pathways involved with receptor tyrosine kinase signalling (R-HSA-1250342 and R-HSA-1963642), and biological processes involved with synapse organisation and signalling (GO:0099177 and GO:0050808). In a previous single cell RNAseq analysis using cells from the entorhinal cortex of AD patients, the *NRG3-ERBB4* ligand-receptor pair was identified to be important for intercellular communication between astrocytes, neurons, oligodendrocyte precursor cells, and other cells. Ablation of *NRG3* and *ERBB4* caused a reduction in excitatory synapse formation in AD patients when compared with healthy controls and affected intercellular communication^38^. *ERBB4* was not genome wide significant in our analysis but achieved nominal significance (*P-value = 2.59×10^−10^*). Additionally, given the association of *NRG3* with cognitive impairment, a hypothesis driven study tested its association with risk and age at onset of AD. The authors identified multiple SNPs and haplotype pairs to be significantly associated with the phenotypes^39^. Hence, despite not being genome wide significant in previous studies, the *NRG3* gene and associated biological pathways have been shown to possess a link to AD.

*PCDH9*, a member of the protocadherin family, facilitates cell adhesion (GO:0007156 and GO:0098742) in neural tissues, contributes to forebrain development (GO:0030900), and is associated or localised in the distal axon (GO:0150034). Variants proximal to or within the gene have been nominally associated with AD^40^, and associated with essential tremor (another neurodegenerative disease)^41^ in previous GWAS. Another GWANN hit, *SMYD3*, has been shown to be significantly elevated in the prefrontal cortex of AD patients and in mouse models of tauopathy. Inhibiting its expression aided in rescuing cognitive defects and restored synaptic function in pyramidal neurons^42^.

#### 3.2 Selection of the significance threshold

We ran the method 16 times to obtain a more stable metric than what would be achieved by running the method a single time. To empirically determine the P-value threshold of significance, we selected pairs of *k* runs—*k in {2, 3, 4, 5, 6, 7, 8}*—from within the 16 total runs, and assessed the stability of the identified hits at different thresholds of P-values (Supplementary Figure 2a, STAR Methods). We defined the stability of hits as the percentage of intersection between a pair of *k* runs. Since the maximum value of *k* for creating paired runs was 8, given a maximum of 16 runs, the empirical P-value threshold was selected as the largest value that ensured 95% stability of hits for 8 runs, instead of 16. We noticed that the significance threshold becomes larger as the number of runs increases (Supplementary Figure 2b). Hence, the threshold for 8 runs would ensure a minimum stability of 95% for hits identified after 16 runs.

While this approach reduced the false positives, it also had the effect of increasing the false negatives. Given the increasing trend in the P-value threshold with number of runs, if the threshold was increased to *1×10^−15^*, three well known AD genes, *PICALM* (*P-value = 6.99×10^−21^*), *EPHA1* (*P-value = 1.7×10^−20^*), and *ABCA7* (*P-value = 1.84×10^−16^*), would be added to the list of GWANN hits. Furthermore, if the P-value threshold was considered as *7.06×10^−7^*, the Bonferroni threshold for multiple testing, instead of empirically determining it, *ACE* (*P-value = 2.39×10^−12^*), *CD2AP* (*P-value = 4.62×10^−12^*), *IL34* (*P-value = 4.52×10^−9^*), and *APP* (*P-value = 6.37×10^−7^*), would also be added to the list of well-known AD hits identified by GWANN. However, for each of the above-mentioned thresholds, 53% and 65% of all the significant genes would have no evidence for association with AD or AD-related traits in previous GWAS. While a proportion of these genes could have true association with family history of AD, others would increase the false positives identified by GWANN and reduce the confidence of the reported hits. Hence, we decided to use the more conservative empirical threshold of *1×10^−25^* to limit the false positives and report the most confident hits.

#### 3.3 Comparison of methods and datasets

We studied the overlap of hit genes and top 100 genes between GWANN, TradGWAS (PLINK 2.0 GWAS on GWANN data), and the EADB GWAS (Figure 3c-d). There was a larger overlap between the hits of (i) GWANN and EADB GWAS (n=5) as compared to the overlap between the hits of (ii) TradGWAS and EADB GWAS (n=2). However, for the top 100 genes, the overlap was the same (n=7) in (i) and (ii). A possible reason for the smaller overlap between the hits in (ii), despite employing similar methods, can potentially be attributed to the lower power in TradGWAS. Additionally, the overlap between the top 100 genes for (iii) TradGWAS and GWANN (n=21) was larger than (i) or (ii). This would suggest that there seems to be a greater effect of similarity in the dataset as compared to the method. Although the EADB GWAS included the signal from the UKB, the inclusion of additional datasets made the signal sufficiently different as compared to the data analysed by TradGWAS and GWANN. The effective dataset used in the EADB GWAS (n=788,989) was almost 10 times the size of the GWANN or TradGWAS data (n=84,220), which also contributed to the power of the analysis. We used multiple approaches to calculate the overlap between the different methods and observed the same pattern (Supplementary Figure 3). While the genes from the EADB GWAS were always the same set of 85 genes, the different methods to get genes from GWANN and TradGWAS were (a) selecting the top 100 genes without LD pruning, (b) selecting the hit gene in an LD block after LD pruning, and (c) selecting the entire LD block after LD pruning and calculating if any gene within this block overlaps with a gene in a different method. The reason we used method (c) was to accommodate the condition where a known AD gene within an LD block would be pruned if another gene in the same block had a higher statistic than it.

#### 3.4 Limitations and considerations

Some well-known AD hits such as *CLU*, *CR1, and TREM2* would not be identified by GWANN even if the P-value threshold would be increased, as discussed earlier. An explanation for missing genes like *TREM2* could be the difference in the minor allele frequencies (MAFs) of the hit SNPs and the SNPs selected for the GWANN analysis. We used a MAF of 0.01 and all SNPs rarer than this were not considered. Furthermore, a caveat of our analysis is the limited genomic region that was analysed. Only SNPs within a gene and in the 2500bp flanking it were considered in the analysis, thereby leaving out most intergenic SNPs. Due to the large computational burden of training a very large number of NNs, we decided to limit the number of SNPs to allow the method to run in a reasonable amount of time. This led to a lot of hit SNPs being excluded from the analysis and could be another possible explanation for not identifying some of the well-known AD hits. We also limited the number of SNPs per NN to a maximum of 50. The number of SNPs per gene ranged from 1-10,000 and this would require modification of the NN architecture due to the wide range of input sizes. The limit of 50 SNPs was imposed to avoid having multiple NN architectures and simplify the analysis. However, this is a limitation of our analysis, and it would be more beneficial to include a much larger range of SNPs to utilise the true potential of NNs in identifying non-linear relationships. Thirdly, the GWANN analysis used a 1:1 ratio of cases:controls to avoid the NNs from overfitting to the majority class. This reduced the sample size as compared to what would be used in a traditional GWAS and possibly influenced the failure to discover genes with weaker signals that would benefit from a larger sample size. Hence, the difference in SNPs and sample size used by GWANN, along with the difference in the model itself as compared to previous GWAS could be the factors contributing to the inability of known AD loci to reach significance in this analysis.

Another limitation of GWANN is the inability to provide SNP level statistics for the hit genes. This makes it difficult to compare GWANN with standard GWAS methods and use GWANN results with packages and tools designed for downstream analyses post GWAS. Packages such as SHAP^43^ and Captum^44^ provide methods such as Shapley additive values and integrated gradients that help in assigning importance to NN input features. However, running the method multiple times to get a more robust metric of performance rendered these methods a lot more complicated to implement due to the non-trivial approach that would be required to combine the importance values across all runs. The NLL of the NNs for each gene serves as an alternative to the effect size estimate that can be obtained from a linear model that is commonly used in GWAS. A gene with smaller NLL suggests stronger association as compared to one with a larger NLL. However, the NLL does not tell us about the direction of effect. Additionally, since we had to run the method more than once, it contributed to increasing the computational cost. Hence, effort is required to make the method scalable and efficient.

Finally, we acknowledge that while the analysed cohort had diagnosed AD cases, the majority were those with family history. Family history has been previously used as a proxy for AD^17,18^, but the findings warrant validation in external cohorts with diagnosed AD cases. While it does not serve as a substitute for external validation, in the absence of it, the series of post-hoc enrichment analyses serve as an additional source of confidence for our findings.

#### 3.5 Conclusion

We applied our method to family history of AD using data from the UKB and introduced a new method to complement the success of existing GWAS methods. GWANN identified genes associated with family history of AD that have previously not been identified by GWAS. A series of post-hoc enrichment analyses provided evidence for differential expression of RNA and proteins associated with the hits between the brains of AD patients and healthy controls. Among the new hits, *LINGO2, NRG3, PALD1, PCDH9, SMYD3,* and *SYNPO* have evidence of association with AD or other neurodegenerative disorders from previous *in-vitro* and *in-vivo* studies. Additionally, enrichment of biological pathways and gene ontologies provided possible explanations for the role of these genes in the processes contributing to AD. Furthermore, *SMYD3, LRRC7, NRG3, PCDH9,* and *ROR1* were identified as tractable targets for drug development. Overall, the findings suggest the potential of GWANN to augment the effort of existing methods in understanding the pathogenesis of AD and other diseases.

### STAR Methods

#### Resource Availability

##### Lead Contact

Further information and requests for resources should be directed to and will be fulfilled or coordinated by the Lead Contact, Upamanyu Ghose.

##### Materials Availability

GWANN results and the summaries for all post-hoc analyses are available in the Supplementary Tables.

##### Data and Code Availability

This research was conducted using data from the UK Biobank Resource under the approved project 15181. Data on brain eQTLS, RNA change in the brain, protein change in the brain, and AD pathology measures were obtained from the Agora AD knowledge portal (https://agora.adknowledgeportal.org), site version 3.3.0, and data version syn13363290-v66. Original code used in the analysis can be found at https://github.com/titoghose/GWANN.

### Method Details

#### Population

We utilised data from the UK Biobank (UKB) (http://www.ukbiobank.ac.uk). The data comprises health, cognitive and genetic data collected from ∼500,000 individuals aged between 37 and 73 years from the United Kingdom at the study baseline (2006–2010)^16,45^. We used imputed SNP genotype data as input to GWANN. UKB genotyping was conducted by Affymetrix using a BiLEVE Axiom array for 49,950 participants and on a further updated using an Affymetrix Axiom array for the remaining 438,427 individuals in this study, based on the first array (95% marker content shared). The released genotyped data contained 805,426 markers on 488,377 individuals. Information on the genotyping process is available on the UKB website (http://www.ukbiobank.ac.uk/scientists-3/genetic-data)^45^. Genotype imputation was performed combining the UK 10K haplotype and Haplotype Reference Consortium (HRC) as reference panels^46^. A number of individuals (*n=856*) either with inconsistencies between their genetic predicted and reported sex, or abnormal number of sex-chromosomes were removed. In addition, 968 outliers were identified based on heterozygosity and missingness, and removed. The dataset was further limited to only individuals of “White British” descent resulting in 409,703 remaining individuals. A genetic relationship matrix along with genome-wide complex trait analysis was used to identify 131,818 individuals with relatives within the dataset, using a relationship threshold of 0.025. Only one person from each pair of related individuals was retained. Only biallelic SNPs with MAF > 1% and imputation quality info score > 0.8 were retained for the analysis, and all indels and multi-allelic SNPs were dropped. For the analysis, we used the imputed genotype dosages.

#### Definition of cases and controls

The cases were defined as individuals with at least one of AD diagnosis (*n=1,176*), or parental history of dementia (*n=40,934*). The parental histories of dementia was defined according to a previous study on family history of AD^17^. Individuals with other neurological disorders^47^ were removed from the control groups. Supplementary Table 6 contains a list of all the neurological disorders along with the UKB fields used to determine the presence of the disorders. We divided the entire range of ages into three groups (age-group1: 38-52, age-group2: 53-61, age-group3: 62-73 years), and paired them with the two possibilities of sex (male and female) to obtain six broad groups—(age-group1, male), (age-group1, female) etc. An equal number of controls (*n=42,110*) were randomly sampled while ensuring that a similar number of cases and controls were included from each of the six broad groups. 80% (*n=67,380*) of the data was used to train the NNs and 20% (*n=16,840*) of the data was reserved as a held-out test to evaluate the performance of the NNs and ascertain association with the phenotype.

#### Neural network model

GWANN follows an architecture with 2 branches that later merge into a single trunk (Figure 1). One of the branches reads contiguous SNPs within a genomic region involving each gene, while the other reads the covariates. The common trunk combines this information to predict family history of AD.

Each sample consists of SNPs and covariates for a homogenous group of 10 cases or controls. The NN was trained to predict if a group was formed by cases or controls. The rationale behind using convolutional layers in our architecture (Figure 1) was to implement “group training”, which allows the NNs of GWANN to consider the group of 10 cases or controls as a single sample, enabling them to identify similar patterns across the individuals in the group. The intuition of this architecture is similar to the concept of retrieval augmented NNs, where the NNs make a prediction using not only a single input sample, but also a candidate set of samples similar to the target sample^48^. The “group training” section is implemented as a 1-dimensional convolution with 32 filters, with the weight filters sliding across the different individuals in the group. This allows the NN to assign a weight to each SNP while being invariant to the different individuals in the group. This is followed by an average-pooling layer which takes the mean of the feature vectors obtained after the convolution operation.

Before passing the output of this section to the densely connected section of the model, they are passed through an “attention” block to focus on important features and ignore features without much information. This block contains a linear layer with *ReLU* activation followed by a *softmax* function that converts the features into probabilities (values between 0 and 1), which are finally elementwise multiplied with the output of the linear layer to weight the features.

The densely connected portion of each branch has 2 blocks with linear layers with *ReLU* activation, having 32 and 16 neurons, followed by batch normalisation, and a dropout probability of 0.5. The final feature vector, obtained from the densely connected portion of the NN focussing on the SNPs, is concatenated with a feature vector (or encoding) generated from the covariates (bottom-left branch of NN in Figure 1) and finally passed through the densely connected end layers of the NN to obtain the final prediction. The covariate encodings are obtained from the penultimate layer of the bottom-left branch. The final prediction block of the NN has 2 blocks with linear layers with *ReLU* activation, having 16 and 8 neurons, followed by batch normalisation, and a dropout probability of 0.1.

Before narrowing down on the described architecture, we tried (i) a multi-layer perceptron architecture without group training or the attention block, and (ii) a branched multi-layer perceptron without group training, with one branch for the SNPs and the second for the covariates. However, we noticed that the models were unable to identify any genes with a significant P-value except *APOE* (Supplementary Figure 4). Hence, we decided to incorporate group training along with the attention block.

#### Training the neural network

Gene locations were mapped according to the Genome Reference Consortium Human Build 37 (GRCh37/hg19). For every gene, SNPs within the gene and in the 2500 bp flanking region (upstream and downstream of the gene) were considered. Since NNs are computationally more intensive than linear models, we set the limit to 2500 bp as a trade-off between increased computational time and including downstream and upstream SNPs in the analysis. This also minimised the chances of overlap between genes which are very close to each other. We divided every gene into windows of maximum 50 SNPs and the final analysis was done on all windows of all genes. A different NN was trained for each window per gene in the entire genome. This resulted in having to train a total of 70,848.

The NNs were trained on a classification task with the objective of minimising NLL. To implement “group training”, a sampler was created to group cases and controls into groups of 10, such that, for an epoch, (i) no individual appeared in more than one group, and (ii) each individual only appeared once within a group. After each epoch, the data sampler shuffled the data to form new groups. Hence, the NNs were not biased to seeing the same groups in every epoch. The only exception to this was in the case of the test or validation sets, where the data was not shuffled, to ensure that the metrics were calculated on the same set of samples for each epoch and for all genes.

We pre-trained the covariate branch (bottom-left branch in Figure 1) and froze the weights while training the NNs for all gene windows. There was no significant difference in performance between freezing the weights of the covariate branch and leaving them as trainable (Supplementary Figure 4). Hence, we employed the former because it provided a speed-up in analysis. The optimiser used to train the NN was Adam, with a learning rate of 5×10^−3^, and batch size of 256. Early stopping, with a patience of 20 epochs, was used during training to avoid overfitting. A validation set of 20% was created from the training set to determine the early stopping epoch.

Finally, due to the weak signal in most gene windows, we noticed that results would vary between multiple runs of the method. Hence, we ran the models 16 times with different random seeds to obtain a more stable metric than what would be achieved by running the model once. The final metric used for each gene window was the 20^th^ percentile of the NLL across all 16 runs. The 20^th^ percentile was used instead of the 80^th^ percentile because a lower NLL is better than a higher one.

The NNs were trained using five NVIDIA GTX 2080Ti and five NVIDIA RTX3090 GPUs. Each GPU was set up to run 4 models in parallel resulting in 40 models running in parallel over all GPUs. PyTorch^49^ version 1.8.1 with CUDA 11.1 was used. A run of the method for 70,848 windows took 55.39 hours.

#### Identifying significantly associated genes

A null distribution of 1000 NLLs, *NLL_null_*, was obtained from a set of NNs trained on the same covariate encodings along with 1000 simulated SNP data generated using the “dummy” command of PLINK 2.0^19^. The dummy data was also trained 16 times using the same random seeds that were used in training the windows of all genes. The metric used for each dummy window was also the 20^th^ percentile of the NLL across all 16 runs. Finally, the P-value of gene window *i* was obtained as *1 – CDF_null_(NLL_i_),* where *CDF_null_* is the cumulative distribution function of the skew normal distribution fit to *NLL_null_*, and *NLL_i_* is the NLL for gene window *i.* Supplementary Figure 5 shows the effect of using different sets of 1000 null NLLS in determining the P-values.

The P-value threshold θ*_1_*, to determine significant association was identified empirically. We split the 16 runs into paired groups of size 8. Along with the empirical P-value threshold θ*_1_*, we set a second threshold θ*_2_=7.06×10^−7^*, the value of the Bonferroni corrected P-value of 0.05 for the number of gene windows tested (*n=70,848*). We then identified the value of θ*_1_* that would ensure that 95% of the significant windows would be significant at a genome wide significance level of θ*_2_* if the method were run another 8 times (Supplementary Figure 2a). In other words, it would ensure 95% stability of the method between two iterations of 8 runs. We repeated the above process for groups of sizes 2, 3, 4, 5, 6, 7 and noticed that θ*_1_* was directly proportion to the size of the groups. Hence, we needed lesser stringent θ*_1_* thresholds to ensure 95% stability, as the number of runs grew larger (Supplementary Figure 2b). Observing this, we finally set θ*_1_=1×10^−25^*, the threshold for 8 runs because this would ensure 95% stability when we identified significant genes using all 16 runs. The true θ*_1_* for 16 runs would possibly be less stringent, but we decided to use this more stringent threshold to minimise the chance of false positives.

#### LD pruning of significant gene windows

After identifying the significant windows within different genes, we calculated the LD between the SNPs within these windows using LDLink^50^. SNPs with r^2^ >= 0.8 were considered to be in LD, and in turn, the genes they were mapped to were also considered to be in LD. The gene with the best NLL within a set of genes in LD was considered as the hit gene.

#### Enrichment and post-hoc analyses

We used information from the Agora AD knowledge portal (https://agora.adknowledgeportal.org) to identify genes that have (i) significant eQTLs in the brain; (ii) change in RNA expression in post-mortem AD brains; (iii) change in protein expression in post-mortem AD brains.

STRING v12^17^ was used to perform PPI analysis of the top 100 genes ranked by their test metric. The parameters for the analysis were (i) **Organism analysed:** Homo Sapiens; (ii) **Statistical background set:** Whole genome; (iii) **Active interaction sources:** Textmining, Experiments, Databases, Co⍰expression, Neighbourhood, Gene Fusion, Co⍰occurrence; (iv) **Minimum required interaction score:** Medium confidence (score=0.400); (v) **Max number of interactors to show:** 1st shell: none (query proteins only); 2nd shell: none.

Pathway enrichment was performed using the R package fGSEA^51^. This enrichment was performed using the test metric for all analysed genes. The enrichment was performed for KEGG, Wiki and Reactome pathways, and GO terms present in the canonical pathways of MSigDB v2023.2.Hs^21^. fGSEA was also used to study the enrichment of DEG sets for different AD-related conditions, in different brain regions from two studies—meta-analysis of AD brain transcriptomic data^24^, and transcriptomic analysis between symptomatic AD, asymptomatic AD and controls^25^.

The disease and trait enrichment analysis was performed with the R package disgenet2r, provided by DisGeNET^22^. The parameters for the analysis were (i) **Organism analysed:** Homo Sapiens [9606]; (ii) **Identifier types used:** [SymbolID]; (iii) **Ontologies used:** ‘CTD_human’, ‘UNIPROT’, ‘CLINGEN’, ‘CGI’, ‘ORPHANET’, ‘PSYGENET’, ‘CURATED’, ‘HPO’, ‘INFERRED’, ‘GWASCAT’, ‘GWASDB’, ‘CLINVAR’, ‘BEFREE’; (iv) **Statistical Test Used:** Fisher test with False Discovery Rate (FDR) p-value correction. At the time of running the analysis, the developers of the disgnet2r package released a new paid tool called disgenetplus2r, which ceased the functionality of the old package. Hence, we were unable to perform the disease enrichment for TradGWAS (GWAS run on GWANN data using PLINK 2.0).

Finally, we used TargetDB^26^ to get a picture of the tractability or suitability of the new GWANN hits for intervention by modalities such as small molecules or antibodies. TargetDB^26^ obtain scores for tractability as well as a multi-parameter optimisation score which takes into account structural information, structural druggability, chemistry, biology, disease links, genetic links, literature information and safety information about the target. We further queried the Open Targets Platform^27^ to identify the known drugs and diseases associated with the novel GWANN hits.

#### Quantification and Statistical Analysis

All NNs were run in Python. Stability testing of the NNs and calculation of P-values by comparing against the null distribution were run in R. To fit a skew normal distribution to the NLLs obtained from dummy data, the ‘selm’ function in R was used. Finally, the P-value was calculated using the ‘psn’ function in the ‘sn’ package. When selecting the final NN architecture, comparison between the number of genes discovered by each architecture was performed using a two-sample t-test (‘ttest_ind’) in Python with the Scipy package. In box and whisker plots, the whiskers represent standard deviation. In line plots, the shaded intervals represent standard deviation.

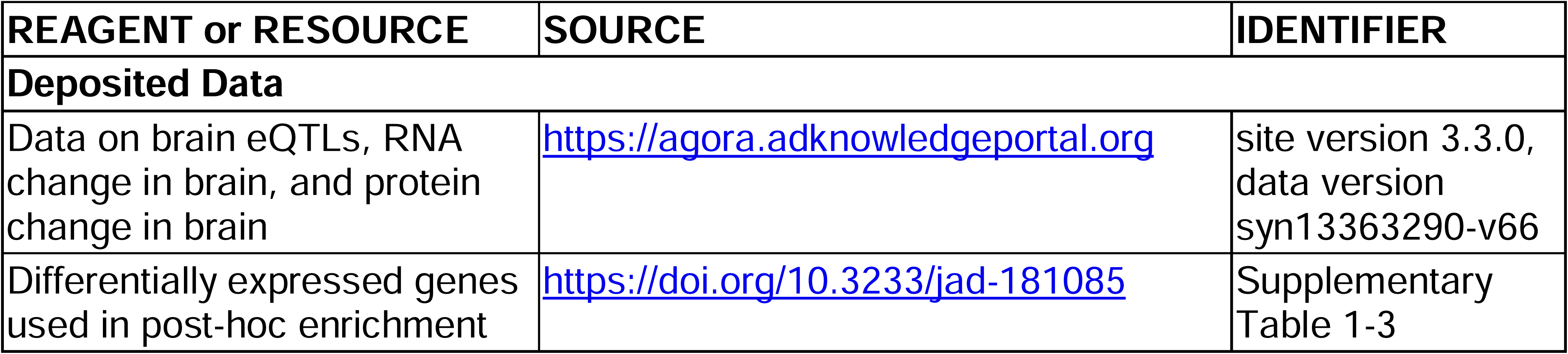
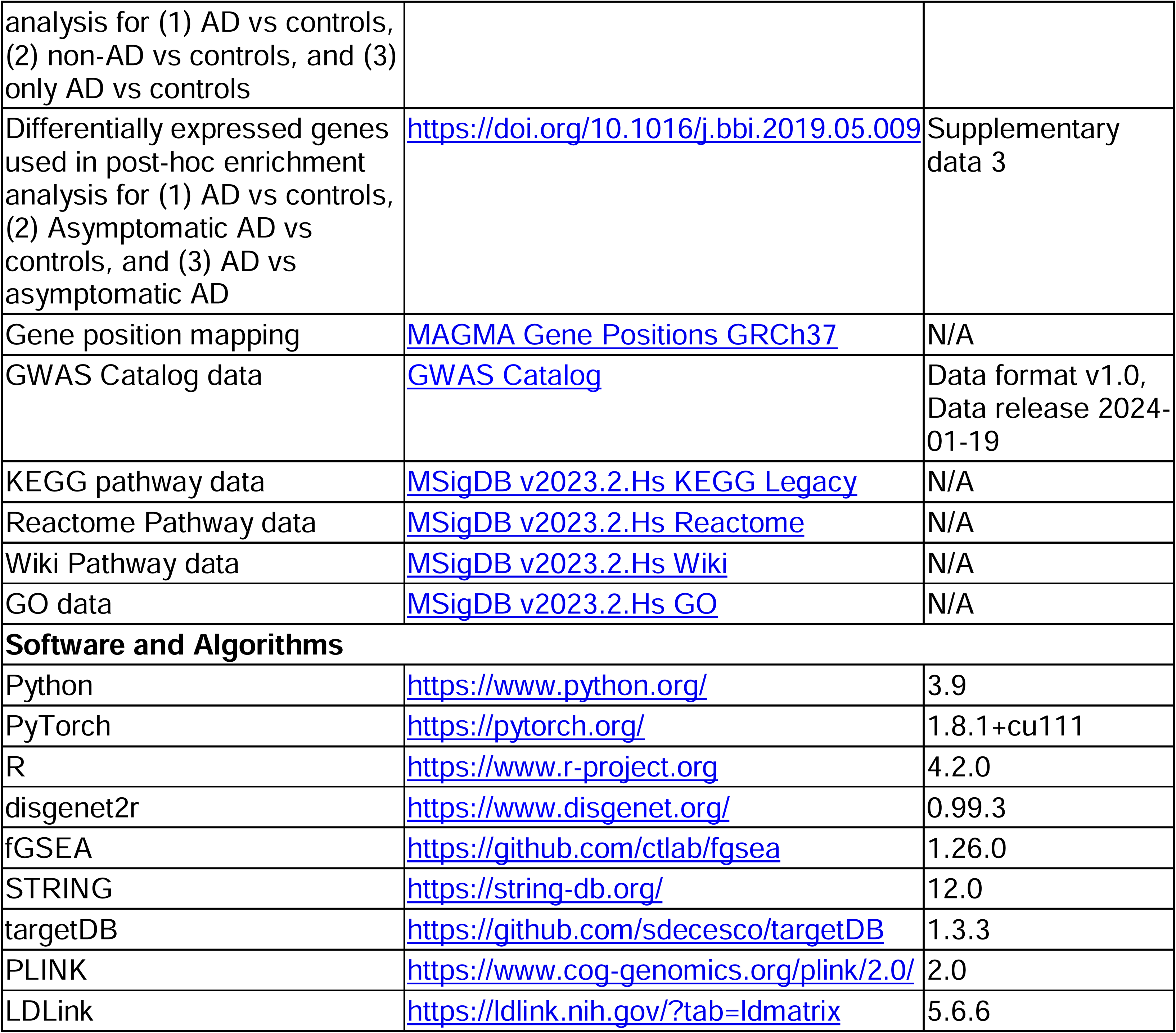
Key Resources Table.

### Supplementary files

**Supplementary Table 1. GWANN summary statistics for all gene windows and all runs.** The columns containing the metric for each run is in the format NLL_x, where x represents the random seed used for the run. NLL and P-value are the aggregate metric and significance values respectively.

**Supplementary Table 2. Results of the GSEA using Reactome, KEGG, Wiki and GO.**

**Supplementary Table 3. Results of the disease and trait enrichment using DisGeNET.**

**Supplementary Table 4. Enriched EFO traits obtained from STRING after constructing the PPI network.**

**Supplementary Table 5. Results of target tractability for drug development using TargetDB.**

**Supplementary Table 6. List of neurological disorders and their associated UKB fields used to filter controls from the GWANN cohort.**

**Supplementary Table 7. Enrichment of DEGs identified in two transcriptomic studies on AD brains using summary stats from the GWAS run on the GWANN data using PLINK 2.0.**

## Data Availability

All data produced in the present work are contained in the manuscript and supplementary information and files.

https://github.com/titoghose/GWANN

## Acknowledgments

We thank the UK Biobank participants and the UK Biobank team for their work in collecting, processing, and disseminating these data for analysis. The results published here are in part based on data obtained from Agora (https://agora.adknowledgeportal.org), a platform initially developed by the NIA-funded AMP-AD consortium that shares evidence in support of AD target discovery.

This work was supported by the Centre for Artificial Intelligence in Precision Medicines (CAIPM); Janssen Research and Development (Johnson & Johnson); the John Fell Foundation [grant ID 0010659]; and the Virtual Brain Cloud from European commission [grant number H2020-SC1-DTH-2018-1]. C.A. is funded by the National Institute for Health Research (NIHR) Oxford Biomedical Research Centre (BRC). The views expressed are those of the author(s) and not necessarily those of the NHS, the NIHR or the Department of Health.

## Author contributions

**U.G.**: Conceptualisation, Methodology, Software, Formal analysis, Visualization, Writing – Original Draft. **W.S.**: Conceptualisation, Methodology, Formal analysis, Writing – Original Draft. **L.W.**: Methodology, Investigation, Writing – Original Draft. **N.A.**: Methodology. **D.N.**: Investigation, Writing – Review & Editing. **B.S.U.**: Writing – Review & Editing. **T.Z.**: Methodology. **A.P.**: Methodology. **Q.L.**: Software. **M.F.**: Formal analysis, Visualisation. **L.S., C.A., A.A., M.A., H.C.**: Writing – Review & Editing. **C.V.D.**: Supervision, Writing – Review & Editing. **A.N.H.**: Conceptualisation, Methodology, Supervision, Writing – Review & Editing.

## Inclusion and diversity statement

We support inclusive, diverse, and equitable conduct of research.

