## Supplementary Material for "Genome wide association neural networks (GWANN) identify genes linked to family history of Alzheimer’s disease"

### Supplementary figures

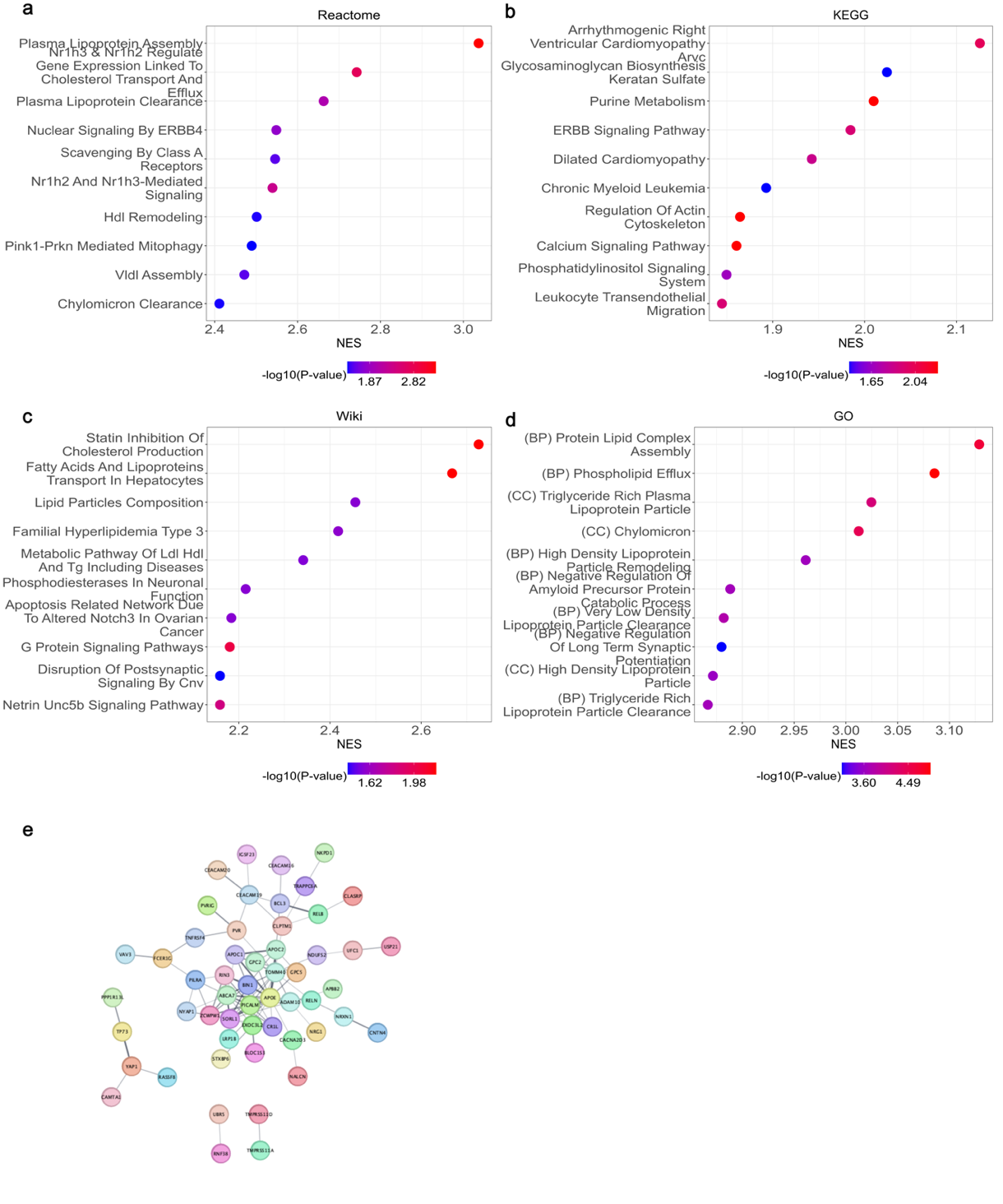

**Supplementary Figure 1.** **Post-hoc enrichment analysis after GWAS on GWANN data using PLINK 2.0.** (a-d) Gene set enrichment analysis for Reactome (a), KEGG (b), Wiki (c), and GO (d) using GWAS summary statistics. (e-f) Genes were ranked according to the absolute z-statistic. (e) Enriched protein-protein interaction network (P-value < 1 x 10^-16^) for the top 100 genes.

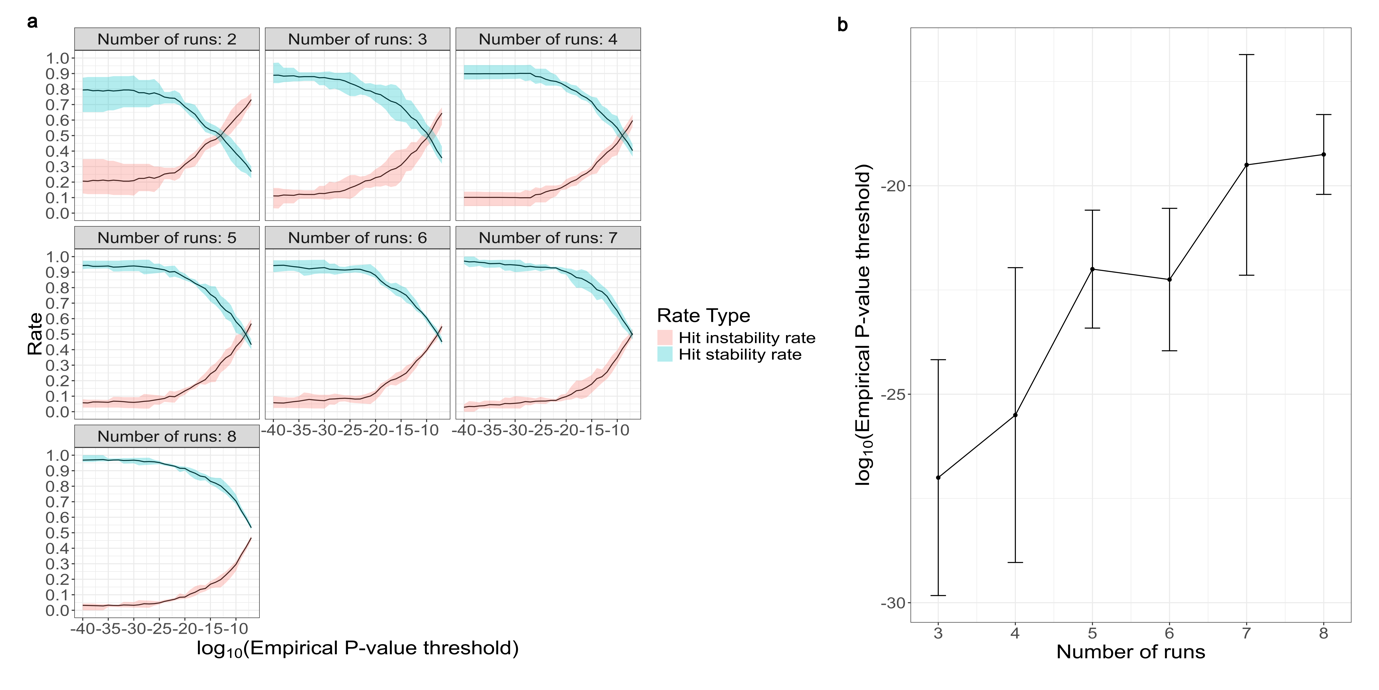

**Supplementary Figure 2. Selection the threshold of significance empirically.** (a) Hit stability and instability rate for different number of runs using different random seeds. The x-axis represents the empirical threshold used to identify significant genes. The stability rate is defined as the ratio of hits that remain significant at the Bonferroni corrected threshold for the number of tests (P-value = 7.06x10^-7^) if the method is repeated a second time for the same number of runs using different random seeds. The instability rate is (1 – hit stability rate). (b) Relationship between the empirical threshold of significance needed to achieve a stability of 90% and the number of runs of the method.

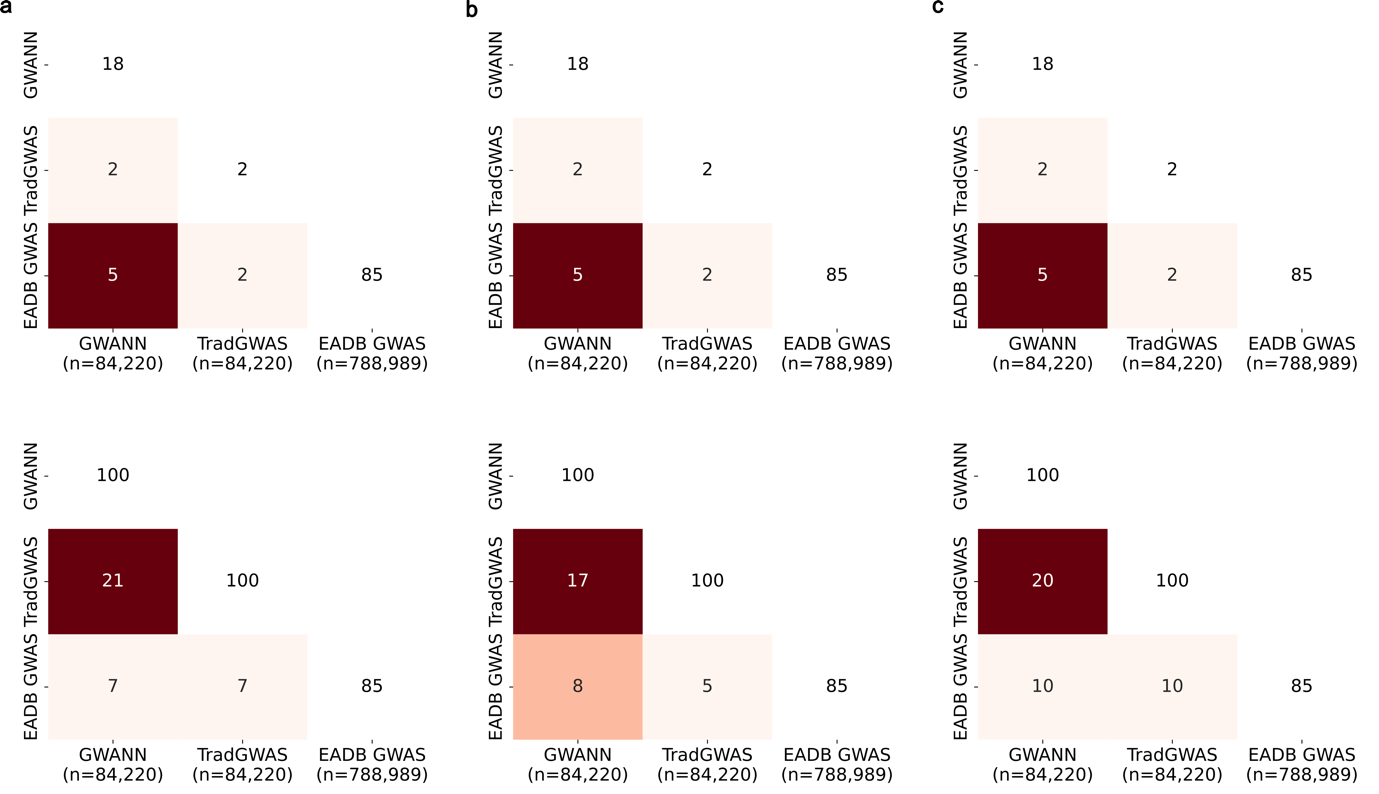

**Supplementary Figure 3. Overlap between GWANN, TradGWAS and EADB GWAS.** (a) Overlap calculated using top 100 genes without LD pruning from GWANN and TradGWAS. (b) The top 100 genes selected from GWANN and TradGWAS were obtained after LD pruning and selecting the gene with the best statistic in an LD block. (c) Instead of selecting a hit gene from an LD block, the top 100 LD blocks were selected while calculating the overlap between methods. The *n* in parenthesis under each method shows the sample size.

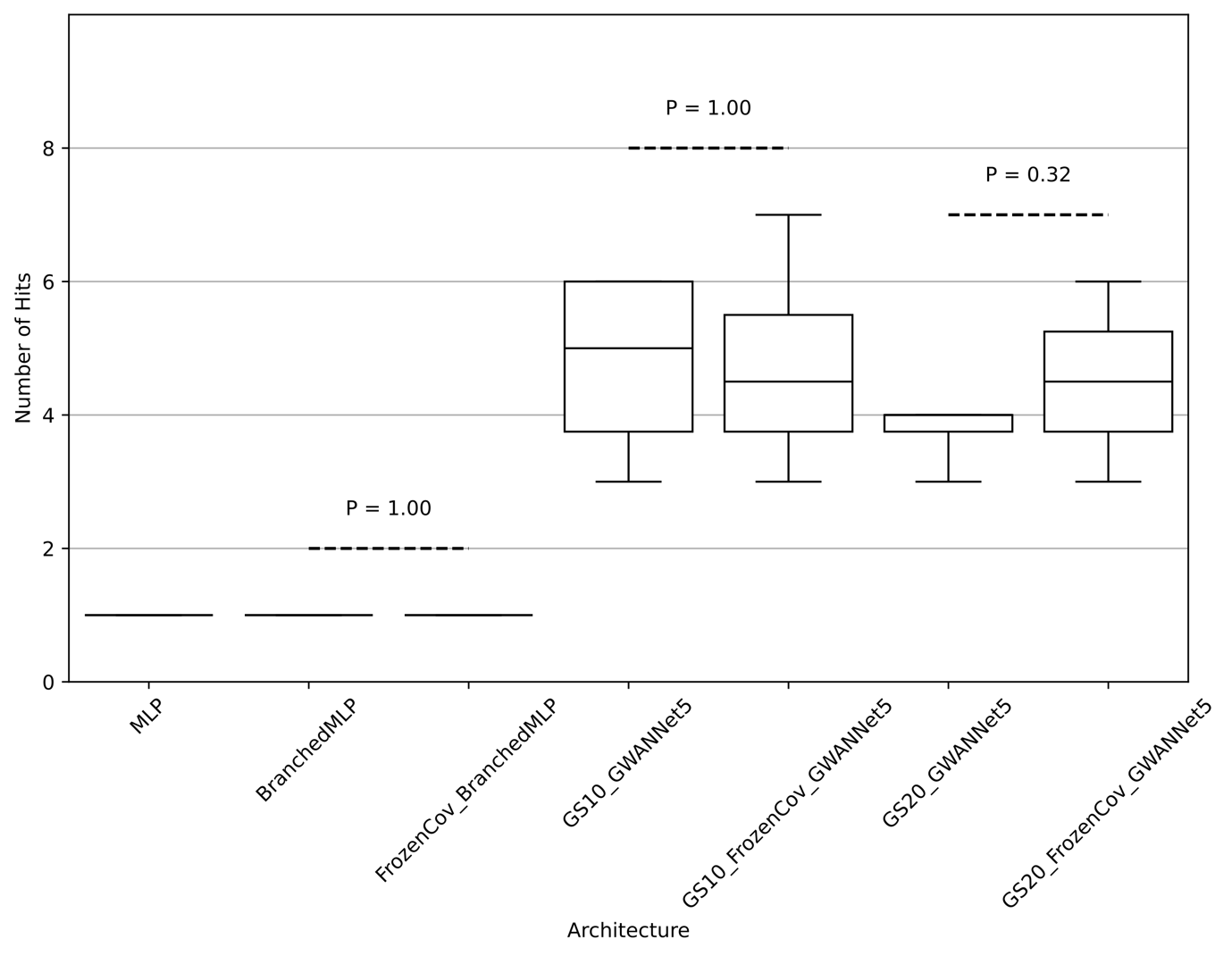

**Supplementary Figure 4. Number of hits identified by different NN architectures from a subset of eight genes.** The tested genes were *APOE, PICALM, BIN1, ADAM10, APH1B, MACROD2, LRRC7,* and *WWOX*. FrozenCov denotes that the covariate branch of the NN (where applicable) was pre-trained and then frozen while training the windows of the genes. Architectures without group training (MLP and BranchedMLP) only identified *APOE* as a hit. For each architecture, we ran the NN three times with different random seeds for each tested gene window and there was no significant difference between freezing the covariate branch and not freezing it.

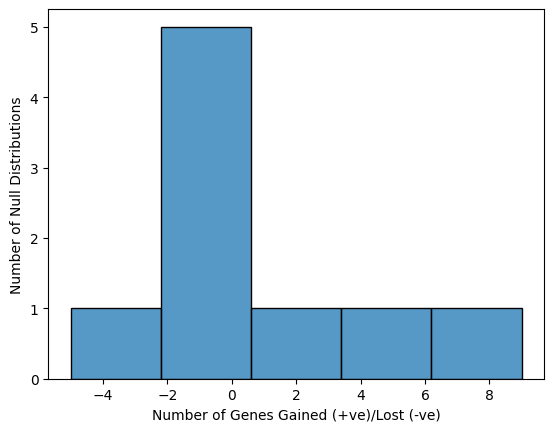

**Supplementary Figure 5. Using different sets of 1000 null NLLs to estimate P-values.** The P-value estimation was run using nine additional null distributions estimated using different sets of 1000 null NLLs in addition to the null distribution used to estimate the P-values reported in the manuscript. The plot shows the change in number of hit genes, compared to the reported hit genes, with each null distribution. Given the stochastic nature of the method, the results showed some variability, but majority of the null distributions showed a change in less than 2 genes from the reported hit genes.
